# Machine Learning–Assisted Feature Selection Identifies the Joint Association of Body Mass Index and Periaortic Adipose Tissue as a Risk Factor for Aortic Dissection: A Multicenter Retrospective Study

**DOI:** 10.64898/2026.04.29.26352087

**Authors:** Shuangjing Wang, Heyue Jia, Pengfei Yuan, Luxia Ren, Mingwei Wu, Haonan Zhang, Peidong Qian, Huan Luo, Yuanming Luo, Zechen Guan, Kai Hou, Min Zhou, Chengkai Hu, Jiang Xiong, Lixin Wang, Weiguo Fu

**Affiliations:** Department of Vascular Surgery, Zhongshan Hospital, Fudan University, Shanghai, China; Institute of Vascular Surgery, Zhongshan Hospital, Fudan University, Shanghai, China; National Clinical Research Center for Interventional Medicine, Shanghai, China; Department of Emergency Surgery, The People’s Hospital of Peking University, Beijing, China; Department of Vascular and Endovascular Surgery, The First Medical Centre, Chinese PLA General Hospital, Beijing, China; Department of Technology, Boea Wisdom (Hangzhou) Network Technology Co., Ltd. Hangzhou, China; Department of Mechanical Engineering, The University of Iowa, Iowa City, USA; Department of Radiology, Zhongshan Hospital, Fudan University, Shanghai, China

**Keywords:** aortic dissection, machine learning, body mass index, periaortic adipose tissue

## Abstract

**BACKGROUND:** Aortic dissection (AD) is a life-threatening emergency with high mortality. Although elevated body mass index (BMI) is associated with both AD incidence and mortality, the underlying mechanisms remain unclear. Periaortic adipose tissue (PAAT) increases with BMI, and the PAAT of AD shows marked inflammatory infiltration, suggesting PAAT-driven inflammation may contribute to the development of AD. However, no direct evidence links BMI and PAAT to AD. To further elucidate the obesity-inflammation-AD relationship, we aim to quantify the contributions of BMI, PAAT, and their derived indices to the risk of AD.

**METHODS:** This retrospective multicenter study (June-November 2025) quantified PAAT around the descending thoracic aorta with CT angiography (CTA). Logistic regression analyses were performed to identify AD risk factors. Based on the Boruta algorithm (a machine learning feature selection method) and ROC curve analysis, the variable importance for AD risk was assessed. The dose–response relationship between BMI–Volume-derived metric (BMV) and AD risk was further characterized by quartile stratification and restricted cubic spline (RCS).

**RESULTS:** This study enrolled 376 consecutive participants. After adjusting for potential confounders, BMI, smoking, systolic blood pressure (SBP), diabetes mellitus (DM), TC/HDLC, ApoE, PAAT volume (Volume), PAAT fat attenuation index (FAI), and BMV were identified as independent predictors of AD. Volume was the strongest AD predictor with the highest Z-score. Compared with BMI [AUC 0.627, 95% confidence interval (CI): 0.569–0.687] and Volume (AUC 0.716, 95% CI: 0.662–0.772), BMV showed better discriminatory performance (AUC 0.726, 95% CI: 0.673–0.778). RCS showed an approximately linear positive association between BMV and AD risk (P-overall < 0.001, P-non-linear = 0.09).

**CONCLUSIONS:** In this retrospective multicenter study, BMV, a composite measure integrating systemic and periaortic adipose tissue factor, showed a positive association with AD risk, and improved predictive performance beyond BMI, indicating incremental predictive value, pending external validation.

**GRAPHIC ABSTRACT:** A graphic abstract is available for this article.

**WHAT IS KNOWN:**

- Body-mass index (BMI) appears to be associated with an increased risk of aortic dissection (AD) and higher all-cause mortality, but a definitive consensus remains elusive.
- PAAT has been identified as an independent cardiovascular risk factor, with marked inflammatory cell infiltration observed in the PAAT of PAAT has been identified as an independent cardiovascular risk factor, with marked inflammatory cell infiltration observed in the PAAT of aortic diseases patients.
- PAAT increases with BMI, and its pro-inflammatory function may destabilize the aortic wall, but relationship of BMI–PAAT synergy and AD remains to be demonstrated.

**WHAT THE STUDY ADDS:**

- BMI, PAAT volume, and its FAI were independent predictors of AD, with volume ranked as the strongest predictor by the Boruta algorithm.
- The BMV, a composite metric that integrated systemic and periaortic adipose tissue factor, was positively associated with AD risk and improved predictive performance beyond BMI alone.

## Introduction

An aortic dissection (AD) is characterized by an intimal tear that allows blood to propagate within the medial layer and create a false lumen.^1^ The pooled incidence of AD was 3.6 to 6.1 per 100,000 person-years with individual studies ranging from 2.0 to 15.0 per 100,000 person-years.^2^ Despite continuous advances in open surgical techniques, endovascular repair, and perioperative management, AD remains a catastrophic event with high mortality.^1^ Therefore, systematically identifying various risk factors combined with rigorous lifelong risk-factor management is essential to reduce both the incidence and the overall burden of AD.^3^

Advanced age, tobacco use, hypertension, and other risk factors are traditionally regarded as the main risk factors for AD, characterized by structural and functional derangements in the aortic intima and media.^1^ According to epidemiological evidence, body mass index (BMI) is not only significantly associated with the incidence of AD, but also with higher all-cause mortality.^4-6^ Periaortic adipose tissue (PAAT) increases with BMI.^7^ Moreover, histopathological analysis of human aneurysm specimens has revealed marked inflammatory infiltration in PAAT,^8^ and multiple studies have implicated PAAT as an independent cardiovascular risk factor.^7,9^ These observations suggest that PAAT may promote AD through proinflammatory pathways and hence represents a potential therapeutic target.^10,11^ The synergistic effect between PAAT and BMI has not been demonstrated to be related to the development of AD, however. The objective of this study is to systematically evaluate the effect of BMI, PAAT, and their derived indices on AD risk macroscopically.

## Methods

The data that support the findings of this study are available from the corresponding author upon reasonable request. This retrospective observational study consecutively recruited inpatients who underwent aortic CT angiography (CTA) between June and November 2025. Chart review of patients ultimately found to be free of AD showed that aortic CTA was ordered for indications including (1) non-contrast chest CT demonstrating aortic dilatation with wall thickening or a possible intimal flap, prompting CTA for clarification; (2) abrupt onset of tearing chest and back pain prompting exclusion of acute aortic syndrome; and (3) pre-operative mapping of the entire aorta prior to coronary artery bypass grafting, valve surgery, or transcatheter aortic valve replacement to guide procedural planning. Based on a three-center CTA database, patients were stratified into two groups according to whether they were diagnosed with AD or not, and risk factors for AD were evaluated. The study protocol adhered to the ethical guidelines of the 1975 Declaration of Helsinki and received approval from the Human Research Ethics Review Board (S2023-597-01). Written informed consent was obtained from all participants. Complete protocols for PAAT quantification (Figure 1), data collection and statistical analysis were provided in the Supplemental Methods.

**Figure 1.**
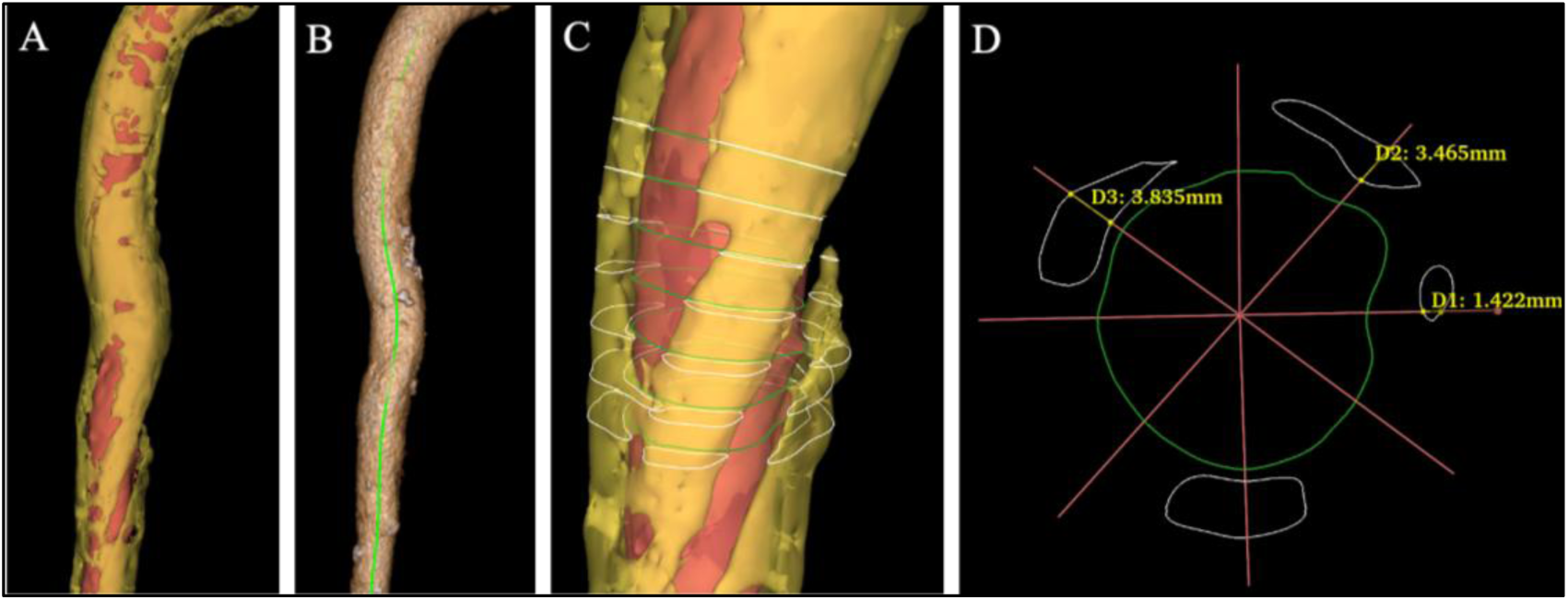
Quantification for PAAT. A,Voxels -190 to -30 HU extracted as PAAT, clipped to 20 mm from wall. B, Three-dimensional thinning, pruning and spline-fitted aortic centerline. C, Minimum Euclidean distance map of adipose voxels to aortic wall. D, 360-ray radial thickness, infiltration angle and circumferential PAAT metrics.

## Results

### Baseline characteristics

This multicentre study was conducted at three tertiary referral centres. Between 1 June 2025 and 30 November 2025, 376 consecutive patients (median age 61 years, 64 % male) who met all eligibility criteria were enrolled (Figure 2). As shown in Table S1, the AD group had a higher male proportion, and higher prevalence of smoking, alcohol consumption, hypertension, CHD, and prior stroke, but a lower prevalence of DM. BMI (24.9 vs 23.6 kg/m², P < 0.001) and SBP were significantly higher in the AD group. Inflammatory and thrombotic markers (WBC, hsCRP, D-dimer, fibrinogen) were markedly increased, whereas GA, HDLC, ApoA-I, and ApoE were significantly reduced. The lipid ratio TC/HDLC was correspondingly less favourable, and Volume, Area, and FAI were all significantly larger (all P < 0.05). Notably, among the derived indices, BMV was higher (137.14 vs 99.30, P < 0.001) and BMA was lower (4.58 vs 4.64, P = 0.017).

**Figure 2.**
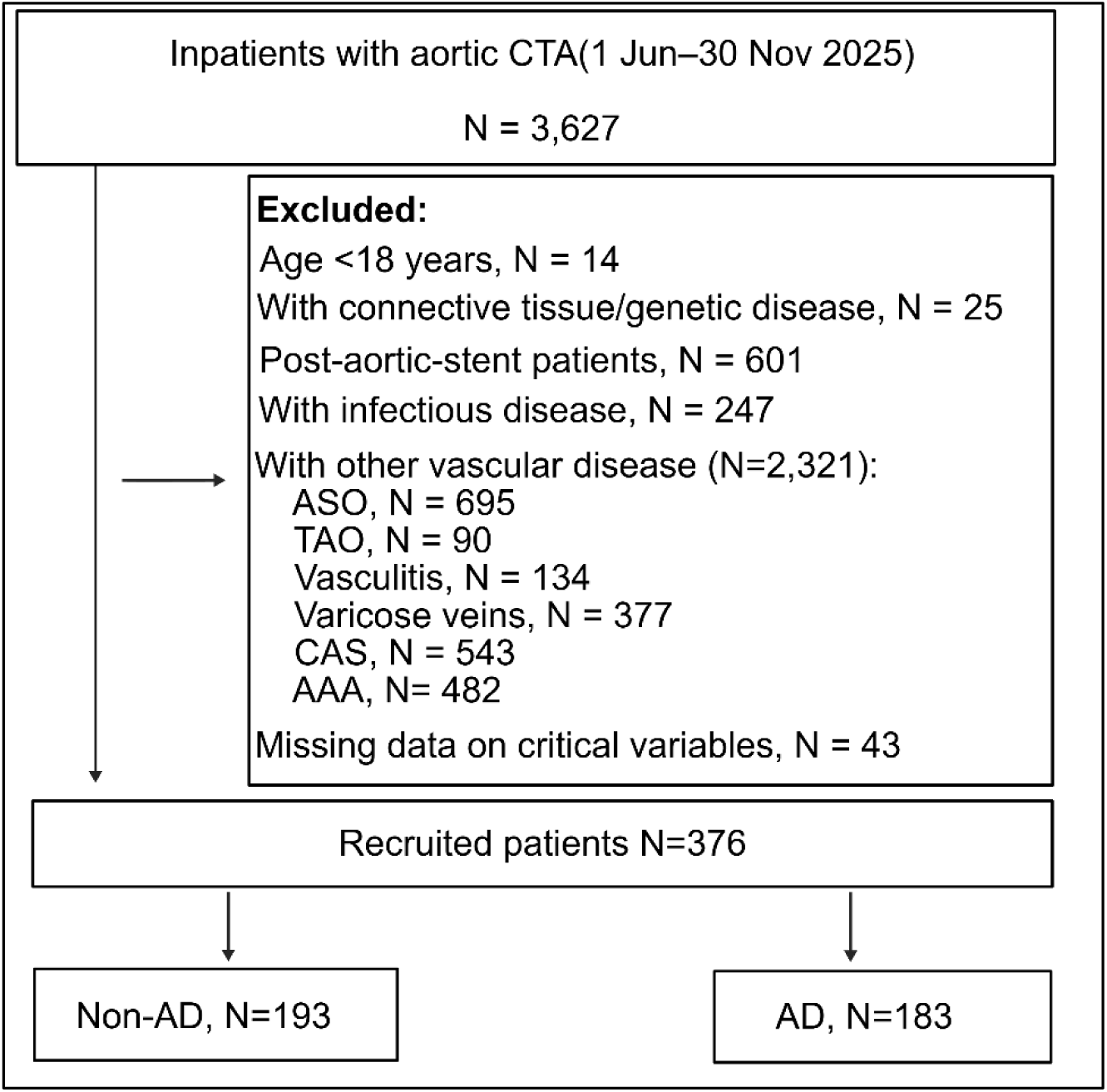
Patient enrollment diagram. ASO, lower-limb atherosclerotic occlusion; TAO, thromboangiitis obliterans; CAS, carotid artery stenosis, AAA, abdominal aortic aneurysm.

### Logistic regression analysis

Thirty two candidate predictors of AD were first examined by univariable logistic regression (**Table S2**). Covariates with P < 0.05 and clinical relevance were considered for inclusion in the multivariable model. Since no severe multicollinearity was detected among BMl, Volume, and BMV (variance inflation factor < 5, Figure S1 and Table S3), BMV was included in the final multivariable logistic reqression model. As shown in Table S4, independent predictors of AD included BMI [odds ratio (OR): 1.131; 95% confidence interval (CI): 1.000-1.280; P = 0.049)], smoking (OR, 10.511; 95% CI: 2.367-46.687; P = 0.002), SBP (OR, 1.036; 95% CI: 1.003-1.070; P = 0.034), DM (OR, 0.162; 95% CI: 0.028-0.924; P = 0.040), TC/HDLC (OR, 5.371; 95% CI: 1.039-27.772; P = 0.045), ApoE (OR, 0.858; 95% CI: 0.784-0.939; P = 0.001), Volume (OR, 1.156; 95% CI: 1.063-1.257; P = 0.001), FAI (OR, 1.167; 95% CI: 1.057-1.289; P = 0.002), and BMV (OR, 1.020; 95% CI: 1.014-1.026, P < 0.001).

### Importance of factors in the impact on AD ranked by Boruta algorithm

The Boruta algorithm was used to evaluate the importance of factors affecting AD (Figure 3). After 500 iterations it was determined that the ariables most closely associated with AD risk (in order of Z-value) were Volume, FAI, TC/HDLC, BMV, GA, SBP, hypertension, ApoE, smoking, DBP, male, Area, ApoA-I, BG, BMI, BMA and alcohol consumption. Notably, Volume achieved the highest importance score and emerged as the strongest predictor of AD.

**Figure 3.**
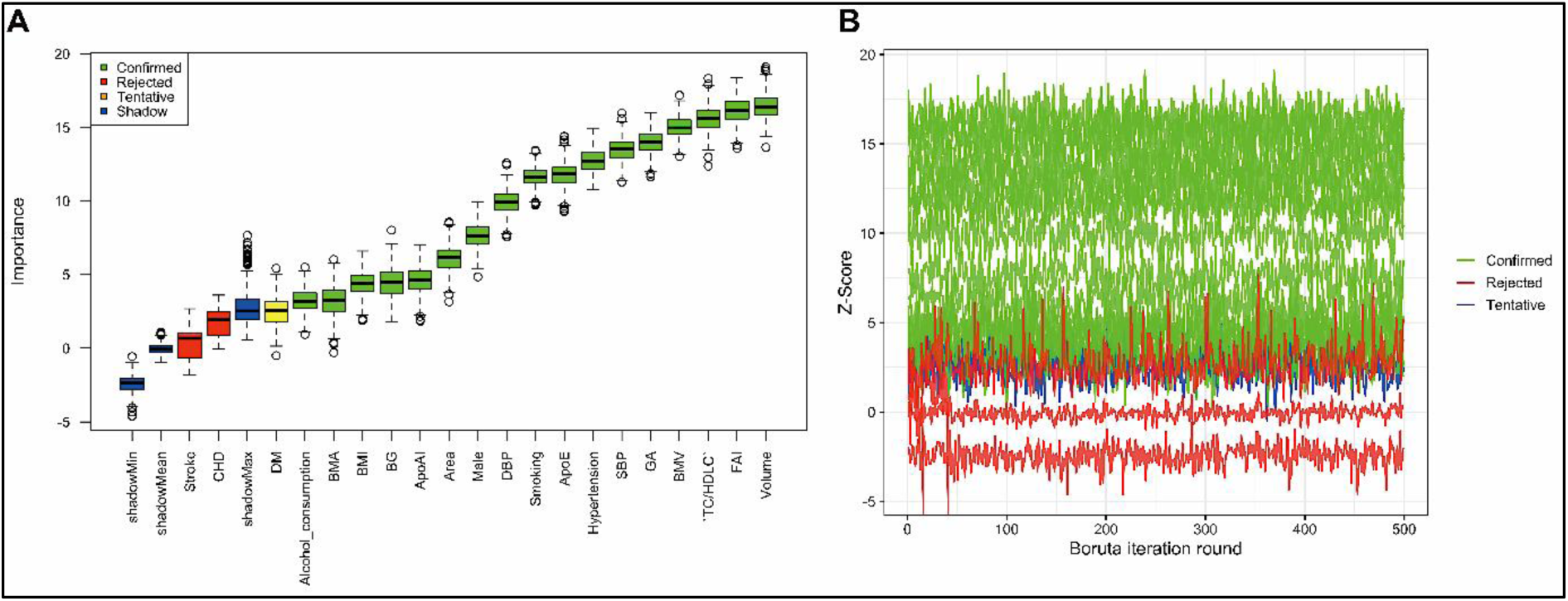
Boruta feature selection between relevant covariates with on AD. A, Importance of factors in the impact on AD ranked by Boruta algorithm. B, Z-Score evolution during Boruta feature selection. SBP, systolic blood pressure; DBP, diastolic blood pressure; DM, diabetes mellitus; BG, blood glucose; GA, glycated albumin; CHD, coronary heart disease; WBC, white blood cell; hsCRP, high-sensitivity C-reactive protein; TC, total cholesterol; HDLC, high-density lipoprotein cholesterol; ApoA-Ⅰ, Apolipoprotein-I; ApoE, apolipoprotein E; FAI, fat attenuation index; BMI, body mass index.

### ROC curves of independent risk factors for AD

For the continuous variables, Volume, FAI, TC/HDLC, BMV, GA, SBP, ApoE, DBP, Area, ApoA-I, BMI, and BG, ROC curve were conducted (Figure 4, Table S5). ROC curves identified four variables with an AUC greater than 0.70, including SBP (AUC, 0.734; 95% CI: 0.683-0.785), BMV (AUC, 0.726; 95% CI: 0.673-0.778), Volume (AUC, 0.716; 95% CI: 0.662-0.772), and DBP (AUC, 0.707; 95% CI: 0.654-0.759). Notably, compared to BMI (AUC 0.627, 95% CI 0.569-0.687), BMV showed superior discriminatory performance (P < 0.001) (Table S6).

**Figure 4.**
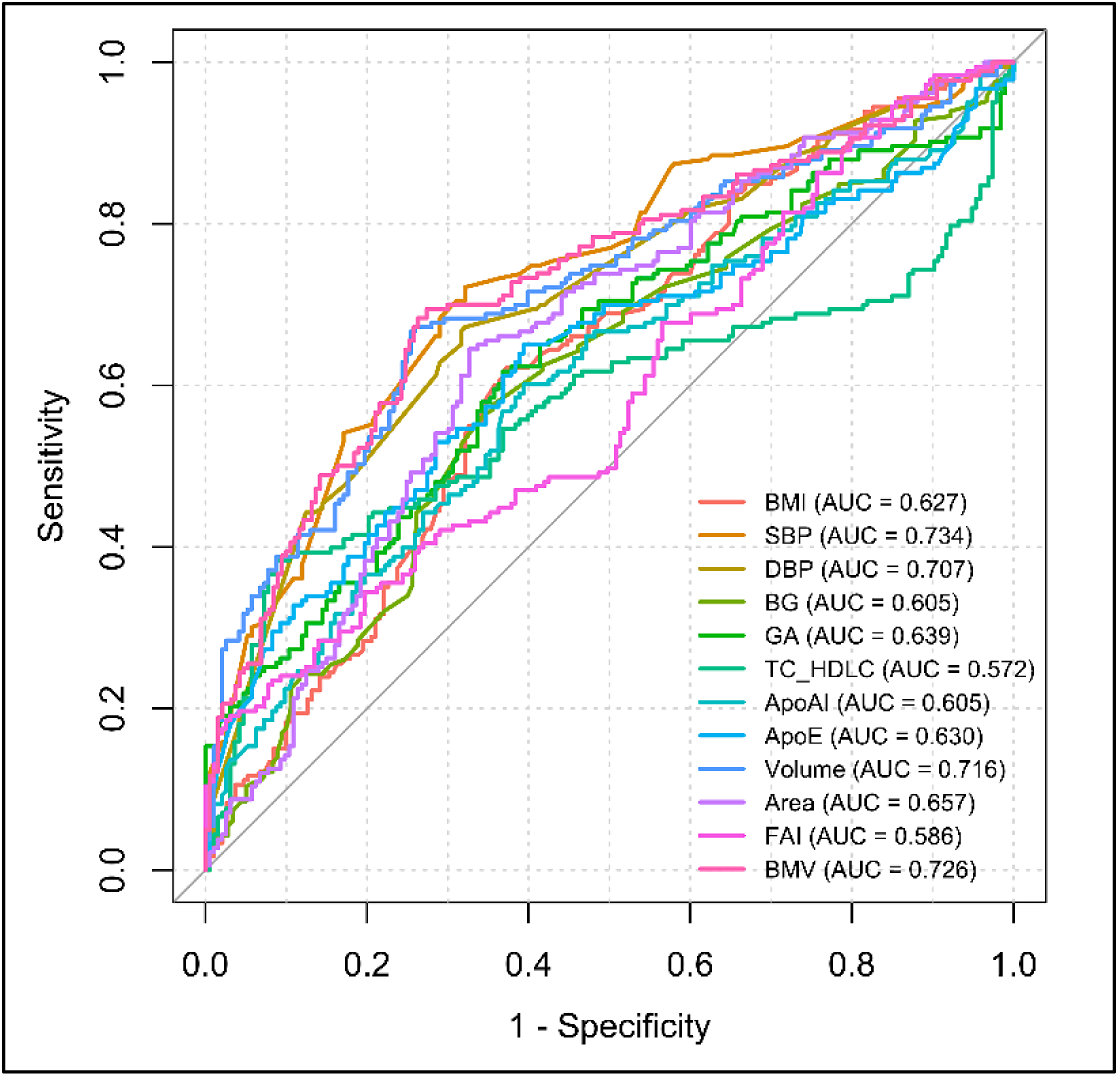
ROC curves for AD prediction. SBP, systolic blood pressure; DBP, diastolic blood pressure; DM, diabetes mellitus; BG, blood glucose; GA, glycated albumin; CHD, coronary heart disease; WBC, white blood cell; hsCRP, high-sensitivity C-reactive protein; TC, total cholesterol; HDLC, high-density lipoprotein cholesterol; ApoA-Ⅰ, Apolipoprotein-I; ApoE, apolipoprotein E; FAI, fat attenuation index; BMI, body mass index.

### Stratification of Patients by BMV Quartiles

Patients were stratified into quartiles based on BMV, including Q1 (BMV < 86.7), Q2 (86.7 ≤ BMV < 114.4), Q3 (114.4 ≤ BMV < 148.7), and Q4 (BMV ≥ 148.7). The characteristics of the four groups were presented in Table 1. The incidence of AD was 29.5% in both Q1 and Q2, then rose to 58.1% in Q3 and 78.5% in Q4. Significant differences were observed across the quartiles for sex, vital signs (SBP, DBP), lifestyle factors (smoking status), comorbidities (hypertension and CHD), laboratory parameters (BG, HbA1c, GA, WBC, Neutrophil, hsCRP, TC, TG, HDLC, TC/HDLC, LDLC, ApoA-I, ApoB, Hcy, Lp(a), ApoE, D-dimer, fibrinogen), PAAT characteristics, BMI, BMA, and BMF (all P < 0.05).

**Table 1.**
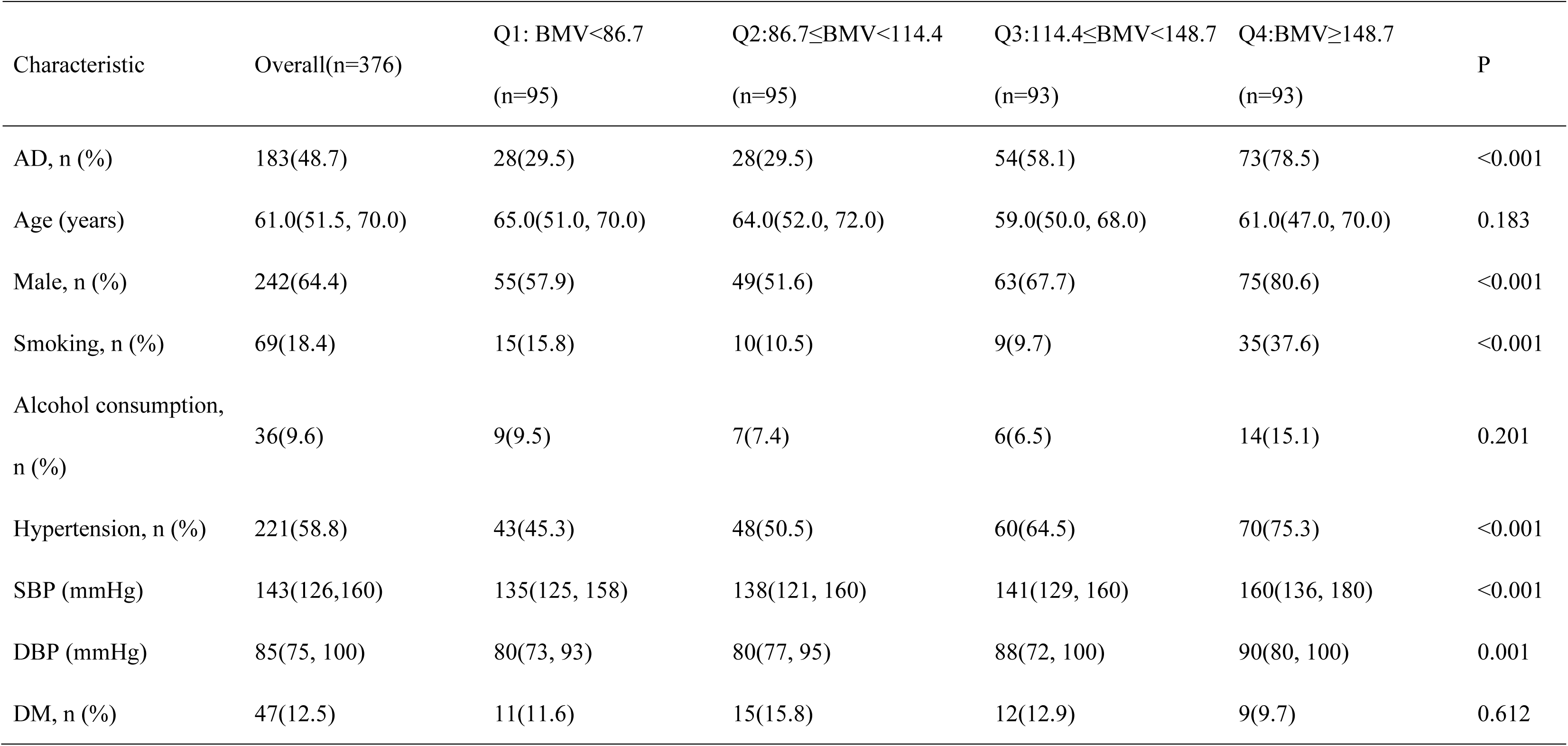
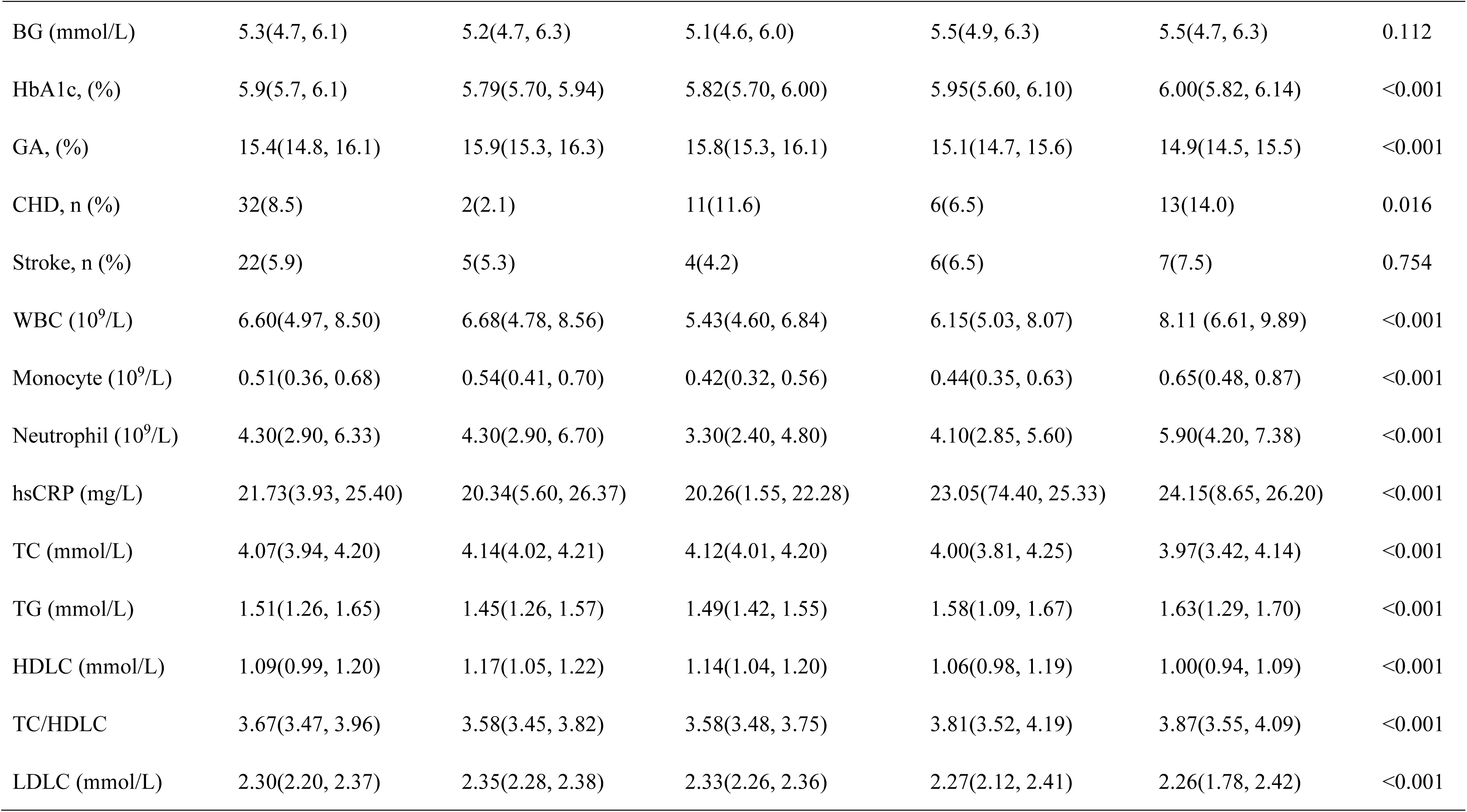
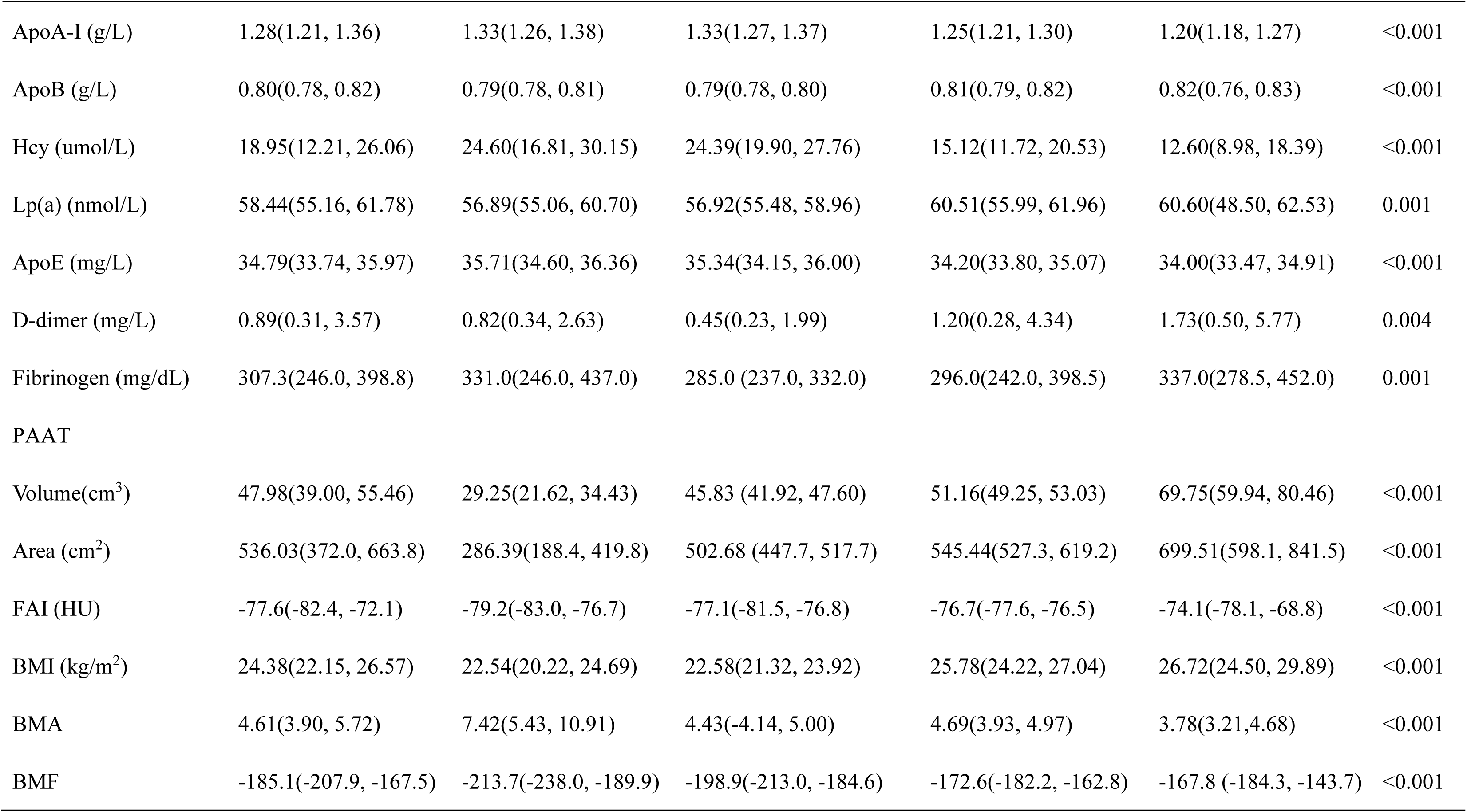
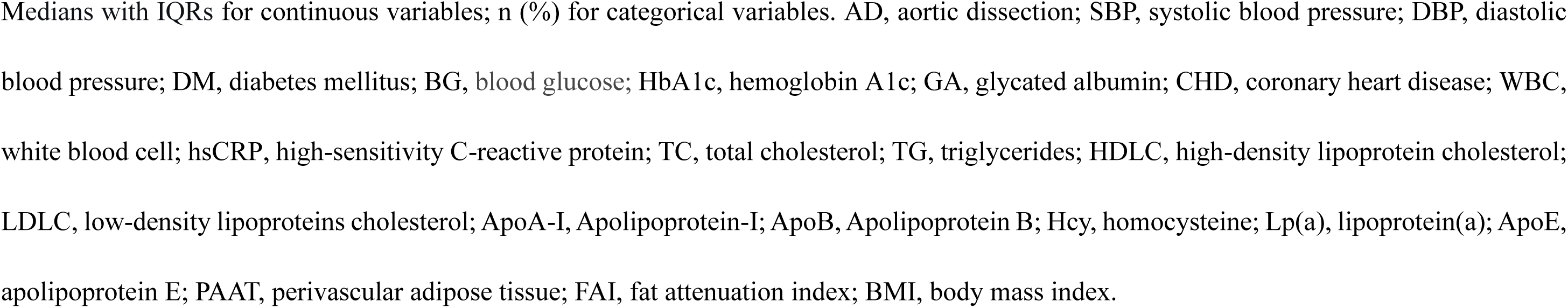
Characteristics of participants categorized by BMV variability.

### Association between BMV and AD

As shown in Table 2, in the fully adjusted Model 3, each unit increase in BMV was associated with an increased risk of AD (OR = 1.046, 95% CI: 1.025–1.067; P < 0.001). Across all adjusted models, the change observed in Q2 did not differ significantly from Q1 (P > 0.05), whereas both Q3 and Q4 were significantly higher than Q1 (P < 0.05). Across all three models, the odds ratios increased progressively in the order Q1 < Q2 < Q3 < Q4.

**Table 2.** Association between BMV quartiles and AD.

| Characteristic | Model 1 |  | Model 2 |  | Model 3 |  |
| --- | --- | --- | --- | --- | --- | --- |
|  | OR (95%CI) | P | OR (95%CI) | P | OR (95%CI) | P |
| BMV | 1.019(1.013-1.024) | <0.001 | 1.015(1.008-1.021) | <0.001 | 1.046(1.025-1.067) | <0.001 |
| Q1: BMV < 86.7 | - | - | - | - | - | - |
| Q2: $86.7 \leq \text{BMV} < 114.4$ | 1.000(0.536-1.866) | 1.000 | 0.981(0.444-2.168) | 0.962 | 1.697(0.558-5.158) | 0.351 |
| Q3: $114.4 \leq \text{BMV} < 148.7$ | 3.313(1.812-6.258) | <0.001 | 2.645(1.243-5.627) | 0.012 | 7.656(1.946-30.125) | 0.004 |
| Q4: $\text{BMV} \geq 148.7$ | 8.734(4.502-16.945) | <0.001 | 5.191(2.346-11.486) | <0.001 | 34.286(4.788-245.497) | <0.001 |

Model 1: No variable adjustment; Model 2: Adjusted for sex, age, DM, CHD, stroke, alcohol consumption, smoking, hypertension; Model 3: Fully adjusted for variables, including sex, age, DM, CHD, stroke, alcohol consumption, smoking, hypertension, BMI, TC/HDL, ApoA-I, ApoE, Volume, Area, FAI. OR, odds ratio; CI, confidence interval.

### Dose–response Relationship

RCS showed that the overall association between BMV level and AD risk was statistically significant (P overall < 0.001) and revealed no significant non-linearity (P nonlinear = 0.09), indicating that the relationship can be regarded as approximately linear, consistent across all three models (Figure 5).

**Figure 5.**
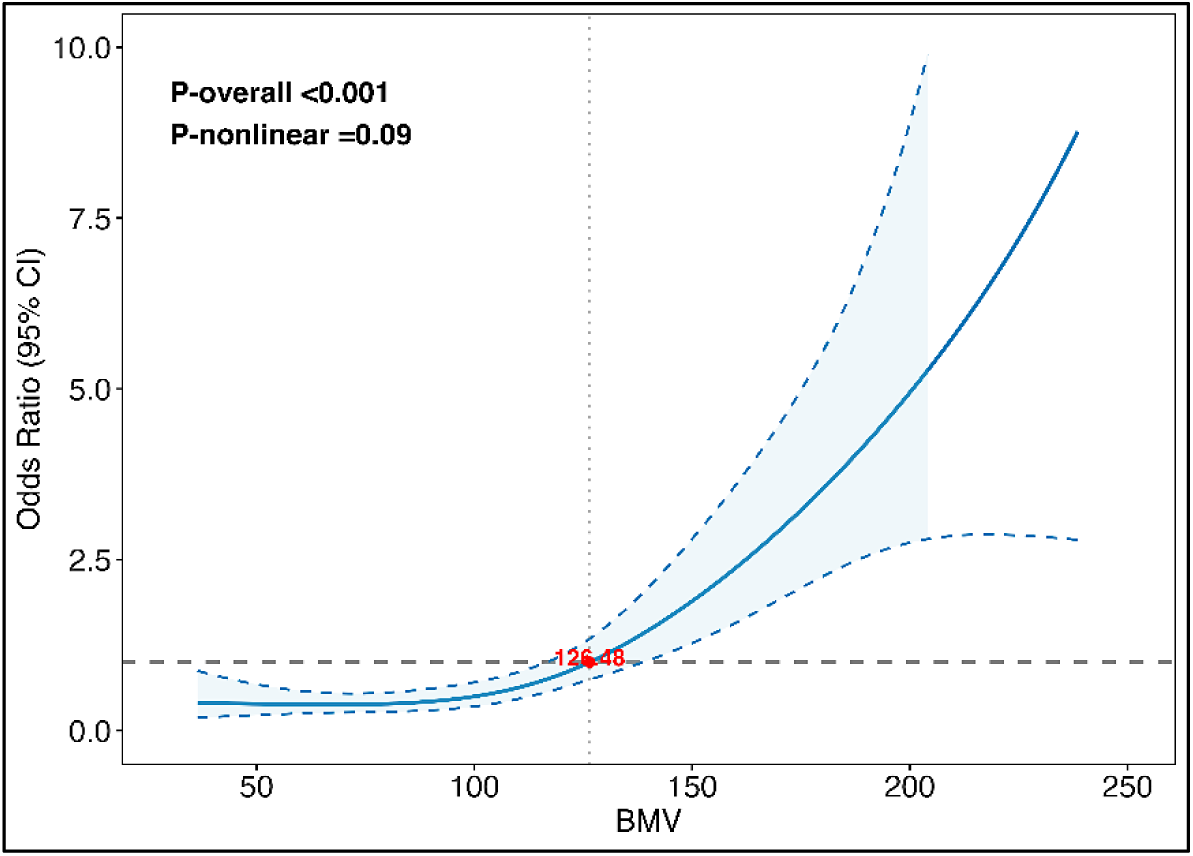
Dose–response relationship between BMV and incident risk of AD.

### Subgroup analysis and interaction tests

Subgroup analyses were performed to assess the relationship between BMV and AD across different patient strata (Figure 6). Except for stroke subgroup, prespecified subgroups (age, sex, CHD, DM, alcohol consumption, smoking status, and hypertension) exhibited a significant positive association between BMV and AD risk (OR > 1, P < 0.05). Within the stroke stratum, the *Yes* subgroup achieved conventional statistical significance (P < 0.001), whereas the *No* subgroup did not (P = 0.0588). Except for hypertension subgroup (P for interaction = 0.033), no significant effect modification was detected in any other subgroup.

**Figure 6.**
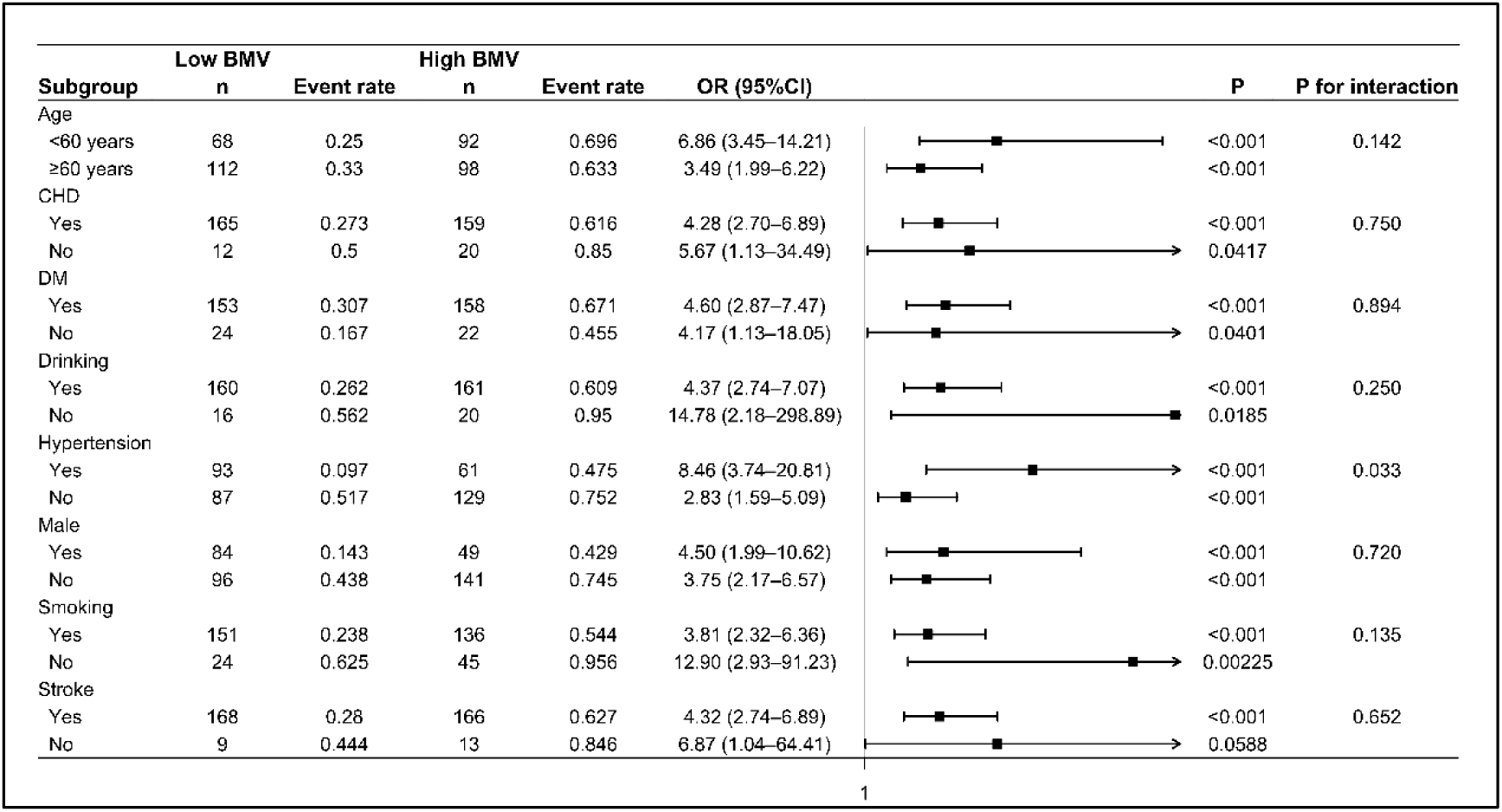
Subgroup analyses of the interaction between BMV and AD. CHD, coronary heart disease; DM, diabetes mellitus.

## Discussion

This multicenter study indicated that systemic and periaortic adiposity, hemodynamic status, and lipid profile were significantly associated with AD. According to multivariate logistic regression, BMI, smoking status, SBP, DM, TC/HDLC, ApoE, Volume, FAI, and BMV were independent predictors of AD. Additionally, BMV exhibited a linear, positive association with AD risk. The ROC curve for BMV yielded an AUC of 0.728 (95 % CI 0.674–0.782). Using the maximum Youden index criterion, the optimal cutoff was 116.967. This threshold remained predictive after adjustment for conventional risk factors and proved robust across major clinical subgroups.

Obesity is well established as a central determinant of traditional cardiovascular events, such as hypertension, CHD, and heart failure,^12,13^ but its influence on AD remains complex and, at times, paradoxical. On the one hand, obesity may act as a precipitating factor, particularly in younger patients.^14^ A nationwide cohort of 2.56 million individuals without prior aortic disease revealed that obesity may potentiate AD among patients with autosomal dominant polycystic kidney disease.^15^ In addition, epidemiological data indicate that BMI is positively associated with both the incidence of thoracic AD and subsequent all-cause mortality.^5,6,16^ On the other, the ‘obesity paradox’ has challenged these observations.^17^ Obesity did not increase mortality or morbidity following surgery for Stanford type A dissection,^18^ and no significant relationship was found between BMI and in-hospital death or post-operative complications after emergent open repair for acute thoracic AD.^19^ Studies leveraging international databases have shown no temporal correlation between BMI and AD mortality in men, and even an inverse linear association in women.^20^ Pathobiologically, the putative link between BMI and aortic disease may be partially mediated by PAAT. Pro-inflammatory mediators released from PAAT can accelerate medial degeneration, the histopathological hallmark of AD.^8,21^ Due to the correlation between BMI and PAAT,^7^ the resultant inflammatory milieu may serve as a mechanistic link between obesity and AD. In this study, although Boruta algorithm identifies Volume as the strongest predictor among all candidates in multivariable logistic regression, ROC analysis indicates BMV achieved a higher AUC than either BMI or Volume, indicating incremental predictive value in capturing AD risk.

In mice, PAAT envelops aorta with asymmetric tri-columnar architecture, leaving the vessel incompletely encased and lacking a continuous circumferential sheath.^22^ Systematic descriptions of the macroscopic distribution of human PAAT are still lacking. Previous investigations have generally adopted 5 mm (from the posterior aortic arch to the diaphragm) ^23^or 10 mm (from 5 cm distal to the aortic root along the ascending aorta)^24^ radial distances to calculate PAAT volume. Given the proportion of PAAT relative to aortic diameter in mice, direct application of a 5-10 mm threshold to humans is likely to underestimate true PAAT volume. We extended the radial boundary to 20 mm from the outer aortic wall and the axial range continuously from the distal left subclavian artery to the aortic bifurcation, thus achieving, a full-length, full-coverage quantification of thoraco-abdominal PAAT. Under this expanded definition, both FAI and Volume remained independent predictors of AD, indicating that the broader measurement strategy did not dilute predictive power, rather, it more faithfully captured the patho-mechanical contribution of PAAT.

Adipose tissue acts as an external cushion that provides physical protection to thoracic and abdominal organs and alters their biomechanical response to external forces.^25^ As reported, ex-vivo thoracic aortic rings surrounded by PAAT exhibited a marked reduction in circumferential stress under identical loading, and PAAT lowers aortic stiffness.^26^ Notably, the mechanical effect of PAAT diminishes with age^27^ and becomes dysfunctional from volume expansion in pathological states.^11,28^ Furthermore, adipose tissue composed of larger adipocytes has attenuation values approaching -190 HU, whereas tissue with smaller adipocytes approaches the water-equivalent value of -30 HU.^29^ Inflammatory cytokines released by diseased vessel wallscan diffuse into PAAT, inhibit adipocyte differentiation, and trigger lipolysis, leading to reduced lipid content, increased water fraction, and a leftward shift of CT attenuation toward - 30 HU.^29^ Pericoronary FAI has already been validated to quantify this adipose attenuation and to serve as a molecular sensor of coronary inflammation, independent of traditional obesity indices such as BMI and waist circumference.^30^ In this study, we further suggest that when PAAT undergoes lipid loss and relative collagen accumulation secondary to inflammation, its buffering capacity declines. This contributes directly to stress concentration within the aortic wall and the initiation of AD.

This multicenter study, a three-stage dimension-reduction pipeline was applied: (1) univariate pre-screening, (2) collinearity diagnosis (after centering the interaction term, VIF < 5), and (3) multivariable logistic regression adjusting for age, sex, blood pressure, and other traditional risk factors, followed by Boruta machine-learning confirmation that BMV ranked among the top four key features. ROC showed that BMV reclassified risk better than BMI or Volume alone. RCS demonstrated a linear dose-response relationship, avoiding black-box effects. However, these findings should be interpreted with caution.

Nevertheless, This study has several limitations. This cross-sectional retrospective study can only demonstrate associations between AD and the examined exposures, it is insufficient for causal inference and requires confirmation in prospective cohort studies. Due to the limited sample size, propensity-score matching was not performed, so baseline imbalances were not fully adjusted for, potentially introducing confounding bias that should be addressed through cross-validation or external validation. Furthermore, the lack of a systematic assessment of AD severity prevents stratified analyses, thus limiting in-depth exploration of the relationship between disease heterogeneity and prognosis.

## Conclusion

In this retrospective multicenter study, the BMV index, the composite measure integrating systemic and local adipose tissue factors, showed a positive association with AD risk, and improved predictive performance compared with BMI, indicating incremental predictive value, pending validation in external cohorts.

## Data Availability

The data that support the findings of this study are available from the corresponding author upon reasonable request.

## Non-standard Abbreviations and Acronyms

PAAT: perivascular adipose tissue
RCS: restricted cubic spline modeling
BMV: body-mass-index-volume

## Acknowledgements

The authors appreciate the Department of Technology staff at Boea Wisdom (Hangzhou) Network Technology Co., Ltd. for generously providing the software and technical guidance required for the calculation of aortic PAAT parameters.

## Sources of Funding

This study was supported by the Noncommunicable Chronic Diseases-National Science and Technology Major Project (2024ZD0537800), National Natural Science Foundation of China (grant number: 82270415, and U24A20651), Shanghai Municipal Science and Technology Commission Fund (grant number: 20234Z00120 and 22S31904800), and Shanghai Sailing Program (grant number: 22QA1408600).

## Disclosures

None.

## Supplemental Material

Supplemental Methods

Table S1–S6

Figures S1

## Notes

### Competing Interest Statement

The authors have declared no competing interest.

### Clinical Trial

This is a retrospective study and trial registration is not applicable.

### Author Declarations

The Institutional Review Board of Chinese PLA General Hospital(S2023-597-01).

